# Skin Lesion Classification Using Convolutional Neural Network for Melanoma Recognition

**DOI:** 10.1101/2020.11.24.20238246

**Authors:** Aishwariya Dutta, Md. Kamrul Hasan, Mohiuddin Ahmad

## Abstract

Skin cancer, also known as melanoma, is generally diagnosed visually from the dermoscopic images, which is a tedious and time-consuming task for the dermatologist. Such a visual assessment, via the naked eye for skin cancers, is a challenging and arduous due to different artifacts such as low contrast, various noise, presence of hair, fiber, and air bubbles, *etc*. This article proposes a robust and automatic framework for the Skin Lesion Classification (SLC), where we have integrated image augmentation, Deep Convolutional Neural Network (DCNN), and transfer learning. The proposed framework was trained and tested on publicly available IEEE International Symposium on Biomedical Imaging (ISBI)-2017 dataset. The obtained average area under the receiver operating characteristic curve (AUC), recall, precision, and F1-score are respectively 0.87, 0.73, 0.76, and 0.74 for the SLC. Our experimental studies for lesion classification demonstrate that the proposed approach can successfully distinguish skin cancer with a high degree of accuracy, which has the capability of skin lesion identification for melanoma recognition.

## 1 Introduction

Skin cancer is a common type of cancer that originates in the skin’s epidermis layer by the irregular cells due to ultraviolet radiation exposure [1]. Every fifth person in the United States (US) has a risk of skin cancer in a region under strong sunshine [2]. Among all skin cancer types, melanoma is the nineteenth most frequently cancer, where approximately 3.0 million new cases were identified in 2018. On average, 2, 490 females and 4, 740 males lost their lives due to melanoma in 2019 [3] in the US alone. It is estimated that approximately, in 2020, 1.0 million of newly affected melanoma patients will be diagnosed. An approximated 6, 850 new case of deaths due to melanoma are anticipated in 2020 in the US alone, which will comprise 4, 610 males and 2, 240 females [4]. Skin cancer deaths in Bangladesh reached 0.04 % of total deaths in 2017, which is ranked 182 in the world [5]. However, a precise and robust early recognition is significant as the survival rate was as high as apparently 90 % in advance recognition [6]. Several imaging techniques like dermoscopic image, optical coherence tomography, magnetic resonance imaging, and confocal scanning laser microscopy are currently being used to diagnose skin cancer. Those images are visually inspected by the dermatologists [7], which is often a tedious and time-consuming process. To relieve the dermatologist’s tediousness by improving the preciseness in recognition, Computer-aided Diagnosis (CAD) systems are being used [8]. Currently, CAD is an integral part of the routine health checkup in the clinic, which consists of raw image acquisition, preprocessing, Region of Interest (ROI) & feature extraction, and finally, recognition [8, 9]. The final step, also known as the classification, is a crucial component, which is a challenging task due to the diverse size of the lesion area, texture, skin color, and the presence of various artifacts like reflections, hair, rolling lines, air bubbles, non-uniform vignetting, shadows, and markers [8, 10], which are shown in Fig. 1.

**Fig. 1:**
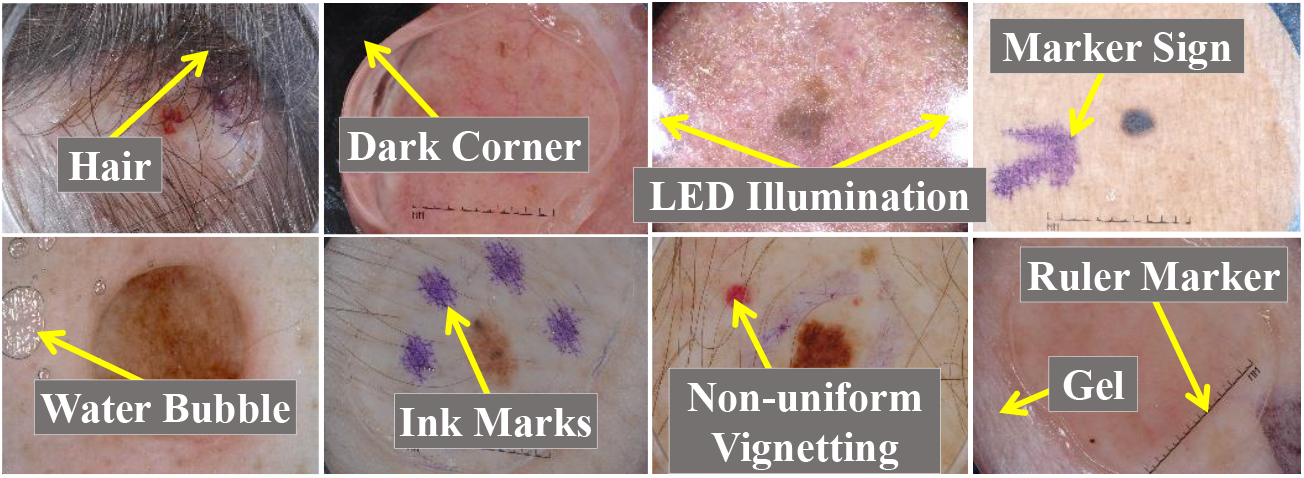
An example of the challenging images, in the ISIC dataset [11], for the accurate SLC.

Nowadays, several methods are being used for the SLC [12, 13]. The sensitivity and specificity of the practitioners are 62 %, and 63 %, while for the dermatologists are 80 % and 60 % respectively for melanoma recognition [14]. In [15], the authors presented a CAD system for the SLC, where they used boarder and wavelet-based texture features with the Support Vector Machine (SVM), Hidden Naïve Bayes (HNB), Logistic Model Tree (LMT), and Random Forest (RF) classifiers. In [16], the authors proposed a model for the SLC, which comprises a Self-generating Neural Network (SGNN), a feature (texture, color, and border) extraction, and an ensembling classifier. A deep residual network (DRN) was presented in [17] for the SLC, where they demonstrated that DRN’s have the capability to learn distinctive features than low-level hand-crafted features or shallower CNN architecture. CNN architecture, along with the transfer learning paradigm, was employed for the SLC in [18]. A 3-D skin lesion reconstruction technique was presented in [19], where depth and 3-D shape features were extracted. Besides, they also extracted regular color, texture, and 2-D shape features. Different machine learning classifiers (SVM and AdaBoost) were employed to classify those features. In [20], the authors proposed an effective iterative learning framework for the SLC, where they designed a sample re-weighting strategy to conserve the effectiveness of accurately annotated hard samples. The stacking ensemble pipeline based on the meta-learning algorithm was proposed in [21], where two-hybrid methods were introduced to combine the mixture classifiers. The effect of dermoscopic image size based on pre-trained CNNs, along with transfer learning, was analyzed in [22], where they resized from 224 × 224 to 450 × 450. They proposed a multi-scale multi-CNN fusion approach using EfficientNetB0, EfficientNetB1, and SeReNeXt-50, where three networks architectures trained on cropped images of different sizes. An architecture search framework was presented in [23] to recognize the melanoma, where the hill-climbing search strategy, along with network morphism operations, were employed to explore the search space.

This study proposes a framework for the SLC (multi-class task), where preprocessing, geometric augmentation, CNN-based classifiers, and transfer learning are the integrated steps. We have performed various types of geometric image augmentations for increasing the training samples as in most of the medical imaging domains, a massive number of manually annotated training images are not yet available [24]. A CNN-based classifier has been used to avoid the tedious feature engineering, which can learn features automatically during the forward-backward pass of training images. Transfer learning of CNN is used to initialize all the kernels in convolutional layers by leveraging the previously trained knowledge rather than random initialization. Extensive experiments are conducted to select different hyper-parameters like types of image augmentation, optimizer & loss function, and metric to be maximized for training, the number of layers of CNN to be frozen. We validate our proposed framework by comparing it with several state-of-the-art methods on the ISBI-2017, where our proposed pipeline achieved better results while being an end-to-end system for the SLC.

The remainder of this paper is set out as follows. Section 2 describes the materials and the proposed framework. Section 3 presents detail results with a proper illustration. Finally, the paper is concluded in section 4 with future works.

## 2 Materials and Methods

The details description of the materials and the methods used in this literature are represented as follows in several subsections:

### 2.1 Dataset and Hardware

The utilized dataset, for training, validation, and testing, is presented in Table 1, where we use the ISIC-2017 dataset from the ISIC archive [11]. Table 1 presents the class-wise distributions and a short description of the ISIC-2017 dataset. The proposed framework was implemented with the python programming language with various python and keras APIs. The experiments were conducted on a *Windows-10* machine with the following hardware configuration: Intel^®^ Core™ i7-7700 HQ CPU @ 2.80 *GHz* processor with Install memory (RAM): 16.0 *GB* and GeForce GTX 1060 GPU with 6 *GB* GDDR5 memory.

**Table 1:** Data distribution of the ISIC-2017 dataset.

| SL # | Class Types | Description | Train | Validation | Test |
| --- | --- | --- | --- | --- | --- |
| 01 | Melanoma (Mel) | Malignant skin tumor obtained from melanocytic | 374 | 30 | 117 |
| 02 | Seborrheic Keratosis (SK) | Benign skin tumor obtained from keratinocytes | 254 | 42 | 90 |
| 03 | Nevus (Nev) | Benign skin tumor obtained from melanocytic | 1372 | 78 | 393 |
| Total Images |  |  | 2000 | 150 | 600 |

### 2.2 Proposed Framework

In the proposed framework, as shown in Fig. 2, the feature extraction and classification of the skin lesion for melanoma recognition have been automated using an end-to-end CNN architecture, where the image augmentation and normalization are the crucial and integral parts of the proposed framework. The deep CNNs are widely used in both medical and natural image classification, which has achieved tremendous success since 2012 [25]. It often rivals human expertise [26]. In CheXNet [27], a CNN was trained on more than 1.0 million frontal-view of the chest X-rays, where it was able to achieve better recognition results than the average recognition by the four experts.

**Fig. 2:**
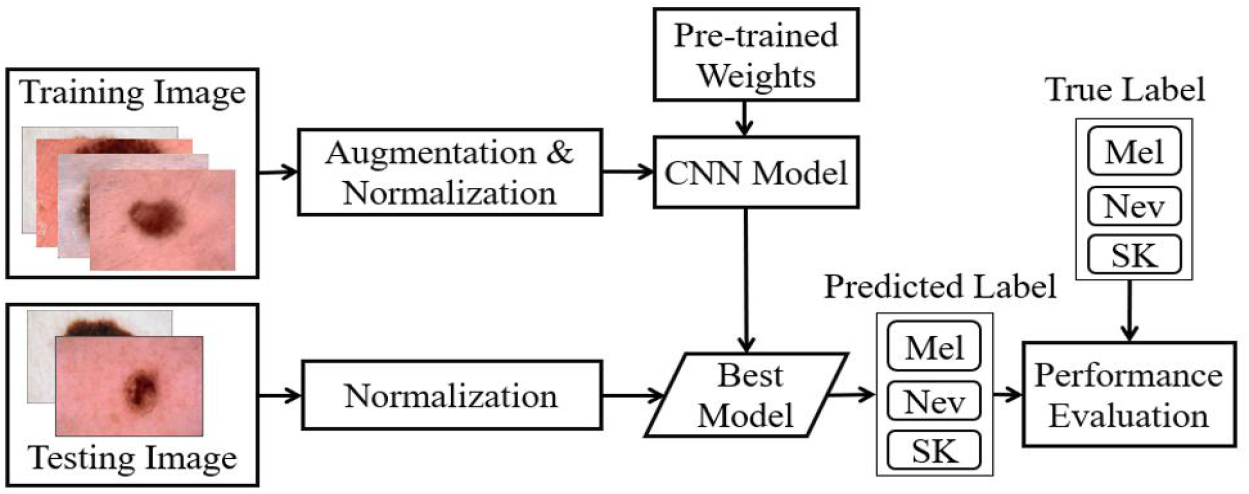
Block diagram of the proposed framework for an automatic SLC towards melanoma recognition using CNN-based classifiers, preprocessing, and transfer learning.

However, in this article, the CNN model is shown in Fig. 3, which has 13 convolutional layers in the 5 convolutional blocks. Each block ends up with a max-pooling layer to lower the computational complexity by reducing the number of connections between convolutional layers, accelerating the CNN models to be generalized by reducing the overfitting. The convolutional layers’ output is fed to the Fully Connected (FC) layers, where we use 3-FC layers and 1-output layer (3 neurons for Mel, SK, and Nev classes). Global average pooling (GAP) was used between convolutional layers and FC layers in place of the traditional flatten layer due to state-of-the-art performance for the image classification [28]. In GAP, only one feature map is generated for each corresponding category, which has an extreme dimensionality reduction to avoid overfitting. In GAP, *height* × *weight* × *depth* dimensional tensor reduced to 1 × 1 × *depth*, where each *height* × *width* feature map transfers to a single number by averaging the *height*.*width* values. Additionally, GAP also provides a lightweight SLC network, which makes it suitable for the real-time SLC-CAD systems for the dermatologists. However, for the fine-tuning the convolutional layers, different layers of feature extractor were frozen to select the optimum number layers to be frozen by maximizing the AUC for the SLC.

**Fig. 3:**
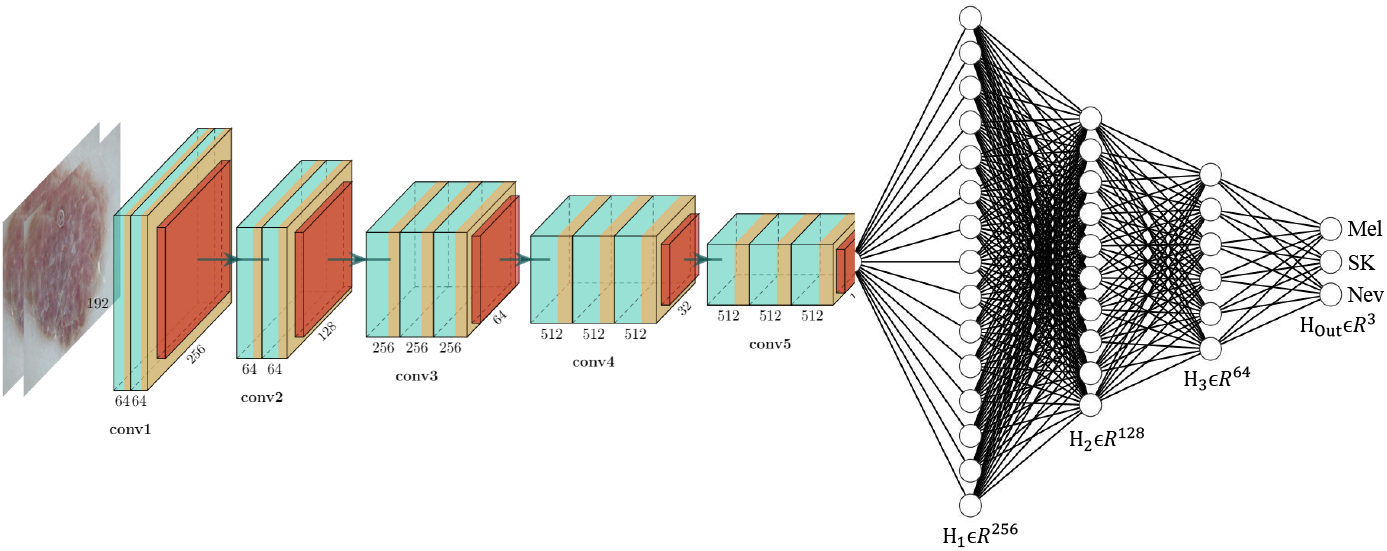
The CNN network for the SLC, where *H*_*m*_ ∈ ℛ^*n*^ is the *m*^*th*^ hidden layer in *n*-dimensional space. The output layer, *H*_*Out*_ lies in 3-dimensional (Mel, SK, and Nev) space.

The scarcity of the training images, especially where the annotation is arduous and costly, can be overcome using a transfer learning [29]. The kernels of the feature extractor (2-D convolutional layers), as shown in Fig. 3, were initialized using the previously trained weights from the ImageNet [30]. The kernels in FC layers were initialized using a glorot uniform distribution [31]. Glorot distribution, also called Xavier distribution, is centered on the mean of 0 with a standard deviation of √(2/(*F*_*in*_ + *F*_*out*_)), where *F*_*in*_ and *F*_*out*_ are the number of input and output units respectively in the weight tensor. Different image augmentation was employed to improve the proposed framework’s generalization ability to the unseen test data, which also makes sure validation error continues to decrease with the training error. Random rotation (0 ∼ 90^*°*^), width shift (10 %), height shift (10 %), random shearing (20 %), zooming (20 %), and horizontal & vertical flipping were used as an image augmentation, in the proposed pipeline, where the outer pixels were filled using a reflection method. The images were also transferred to [0 1] before feeding them into the CNN network. As we saw from the Aspect Ratio (AR) distribution of the ISIC-2017 images, AR lies in 3 : 4. So, we resized the input images to 192 × 256 pixels using the nearest-neighbor interpolation technique [8]. Categorical cross-entropy was the loss function, which was optimized to maximize the average accuracy of lesion classification in our framework. The loss function was optimized using the Adam [32] optimizer with initial Learning Rate (*LR*), exponential decay rates (*β*_1_, *β*_2_) as *LR* = 0.0001, *β*_1_ = 0.9, and *β*_2_ = 0.999 respectively without AMSGrad variant. The initial learning rate was reduced after 5 epochs by 20.0 % if validation loss stops improving. The proposed pipeline was trained in a machine, as mentioned in subsection 2.1, with a batch size of 8.0.

### 2.3 Evaluation Criterion

The proposed pipeline was evaluated using the confusion matrix of True Positive (TP), False Positive (FP), False Negative (FN), and True Negative (TN). The recall, precision, and F1-score were also used, where the recall quantifies the type-II error (the sample with target syndromes, but wrongly fails to be refused) and the precision measure the percentage of correctly classified positive patients from all positive recognition. The F1-score indicates the harmonic mean of recall and precision, which shows the tradeoff between them. Additionally, how well lesion prediction is ranked rather than their absolute value is measured using the AUC.

## 3 Results and Discussion

In this section, the results on the ISIC-2017 test dataset (see in Table 1) for the SLC are presented. At the end of this section, state-of-the-art methods are compared against the proposed framework on the same dataset.

A classification report, to visualize the precision, recall, F1-score, and support scores per-class basis, is shown in Table 2. A classification report is a deeper intuition for the quantitative evaluation of the classifier, which can also show the weaknesses in a particular class of a multi-class problem. The results, as shown in Table 2, show that the correctly classifying samples of Mel, Nev, and SK are 62.0 %, 76.0 %, and 73.0 % respectively, while the respective type-II error (false-negative rate) is 38.0 %, 24.0 %, and 27.0 %. The support-weighted recall, 73.0 % indicates that 27.0 % samples having target symptoms, but erroneously fails to be rejected by the proposed framework. The support-weighted precision, 76.0 % indicates that only 24.0 % samples are wrongly classified among all the recognized true classes. The F1-score reveals that both the precision and recall are better from the proposed framework for the SLC of the ISIC-2017, although the uneven class distribution was being used for training. The more details the class-wise investigation, of the proposed SLC, has been present in the confusion matrix in Table 3. The confusion matrix, as shown in the Table 3, presents the number of correct and incorrect predictions by the proposed framework with a count value. Table 3 shows that among 117, 393, and 92 samples of Mel, Nev, and SK classes are classified as (72, 28, and 17), (57, 300, and 36), and (8, 16, and 66) respectively to Mel, Nev, and SK. Among 117 samples of the Mel classified as the Nev, which is undesirable FN. Table 1 shows that a more number of samples are in the Nev class in the training dataset, which makes the biased classifier towards Nev. The results, as shown in Table 3, also depicts that a significant number of the Mel and SK samples are predicted as Nev. Some of the miss-classified images by the proposed pipeline is shown in Table 4, where we show the difficulties for the correct classifications. From Table 4, it is seen that images (first two rows) having a true class of Mel are classified as SK and Nev with respective confidence probability of 0.934 and 0.88. Similarly, all other images in Table 4 are wrongly classified with a degree of confidence probability. If we visually inspect those images, it seems those images, especially in the fifth row, seem like a Mel as the texture of the lesion area is more complex. However, the dermatologist tells that it is Nev. The probable reason behind such a wrong classification by the proposed framework is the lack of diversity in the training samples, inter-class diversity, and intra-class similarity.

**Table 2:** Classification report for the SLC, where the weights of the classes were calculated from the supported samples for averaging the metrics.

| Class | Precision | Recall | F1-score | Support |
| --- | --- | --- | --- | --- |
| Mel | 0.53 | 0.62 | 0.57 | 117 |
| Nev | 0.87 | 0.76 | 0.81 | 393 |
| SK | 0.55 | 0.73 | 0.63 | 90 |
| Weighted Average | 0.76 | 0.73 | 0.74 | 600 |

**Table 3:**
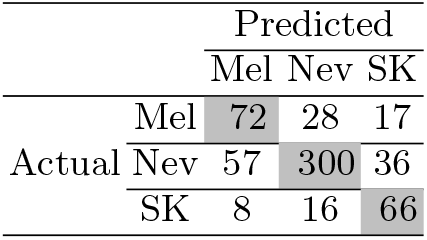
Confusion matrix of the test results, where each column and row represent the instances in a predicted and actual class respectively.

|  |  | Predicted |  |  |
| --- | --- | --- | --- | --- |
|  |  | Mel | Nev | SK |
| Actual | Mel | 72 | 28 | 17 |
|  | Nev | 57 | 300 | 36 |
|  | SK | 8 | 16 | 66 |

**Table 4:**
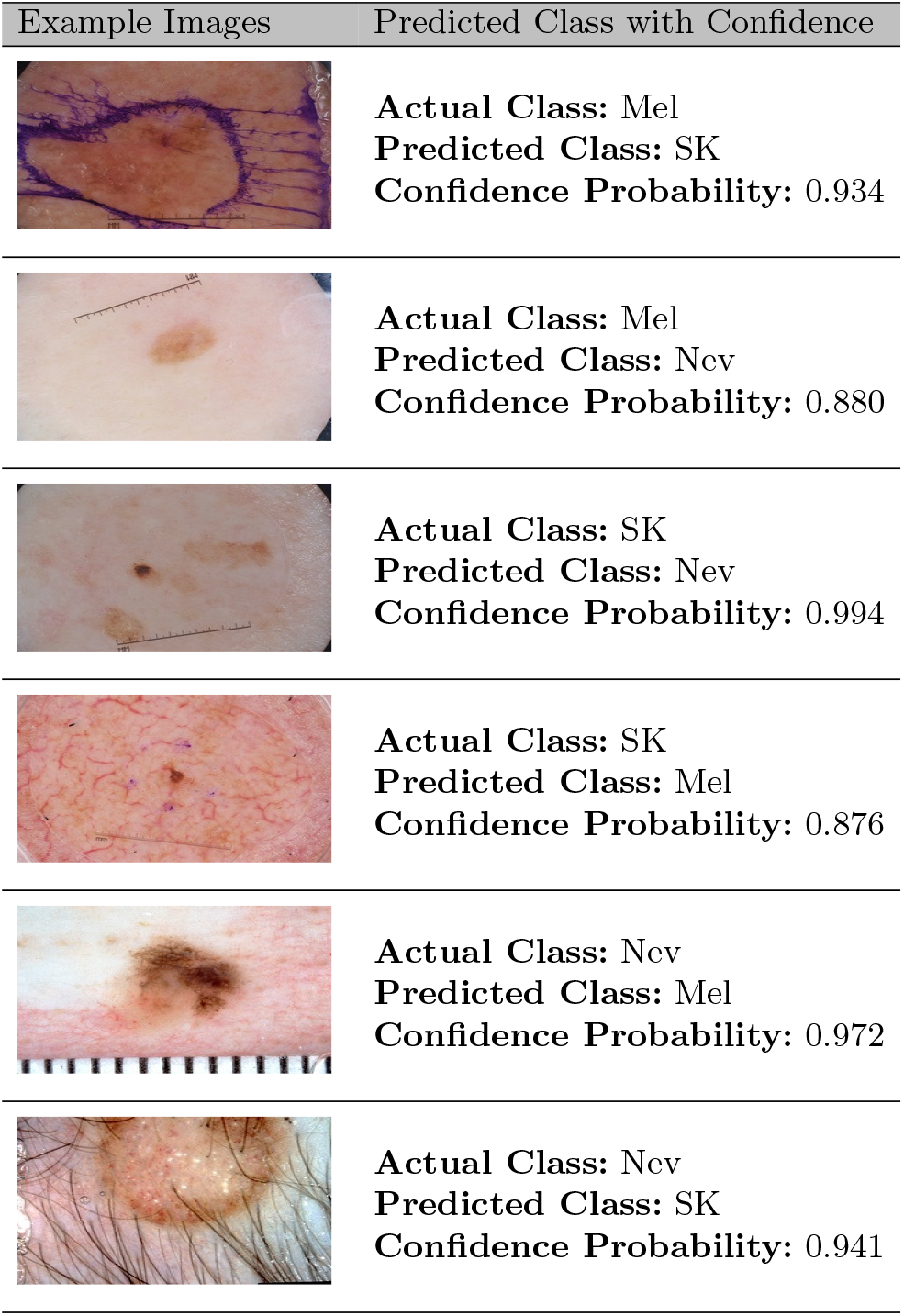
Several examples of wrongly classified images, from the proposed framework, with confidence probability.

| Example Images | Predicted Class with Confidence |
| --- | --- |
|  | **Actual Class:** Mel<br>**Predicted Class:** SK<br>**Confidence Probability:** 0.934 |
|  | **Actual Class:** Mel<br>**Predicted Class:** Nev<br>**Confidence Probability:** 0.880 |
|  | **Actual Class:** SK<br>**Predicted Class:** Nev<br>**Confidence Probability:** 0.994 |
|  | **Actual Class:** SK<br>**Predicted Class:** Mel<br>**Confidence Probability:** 0.876 |
|  | **Actual Class:** Nev<br>**Predicted Class:** Mel<br>**Confidence Probability:** 0.972 |
|  | **Actual Class:** Nev<br>**Predicted Class:** SK<br>**Confidence Probability:** 0.941 |

The ROC and precision-recall curves are shown in Fig. 4 (a) and Fig. 4 (b), respectively. It is observed from Fig. 4 (a) that for a 10.0 % false-positive rate, the true-positive rates for the SLC is approximately 62.0 %. The corresponding macro-average AUC from the ROC curve is 0.873, which indicates that for any given random sample, the probability of accurate recognition as Mel, Nev, and SK is as high as 87.3 %. The precision-recall curve, as shown in Fig. 4 (b), shows the tradeoff between precision and recall for different thresholds, where the high area under the curve represents both high recall and precision. High scores for both show that the proposed pipeline is a blessing with the accurate results as well as returning a majority of all positive results (high recall). The macro-average precision from the proposed pipeline is 80.6 %, which indicates that the proposed pipeline is well-suited for the SLC for melanoma recognition.

**Fig. 4:**
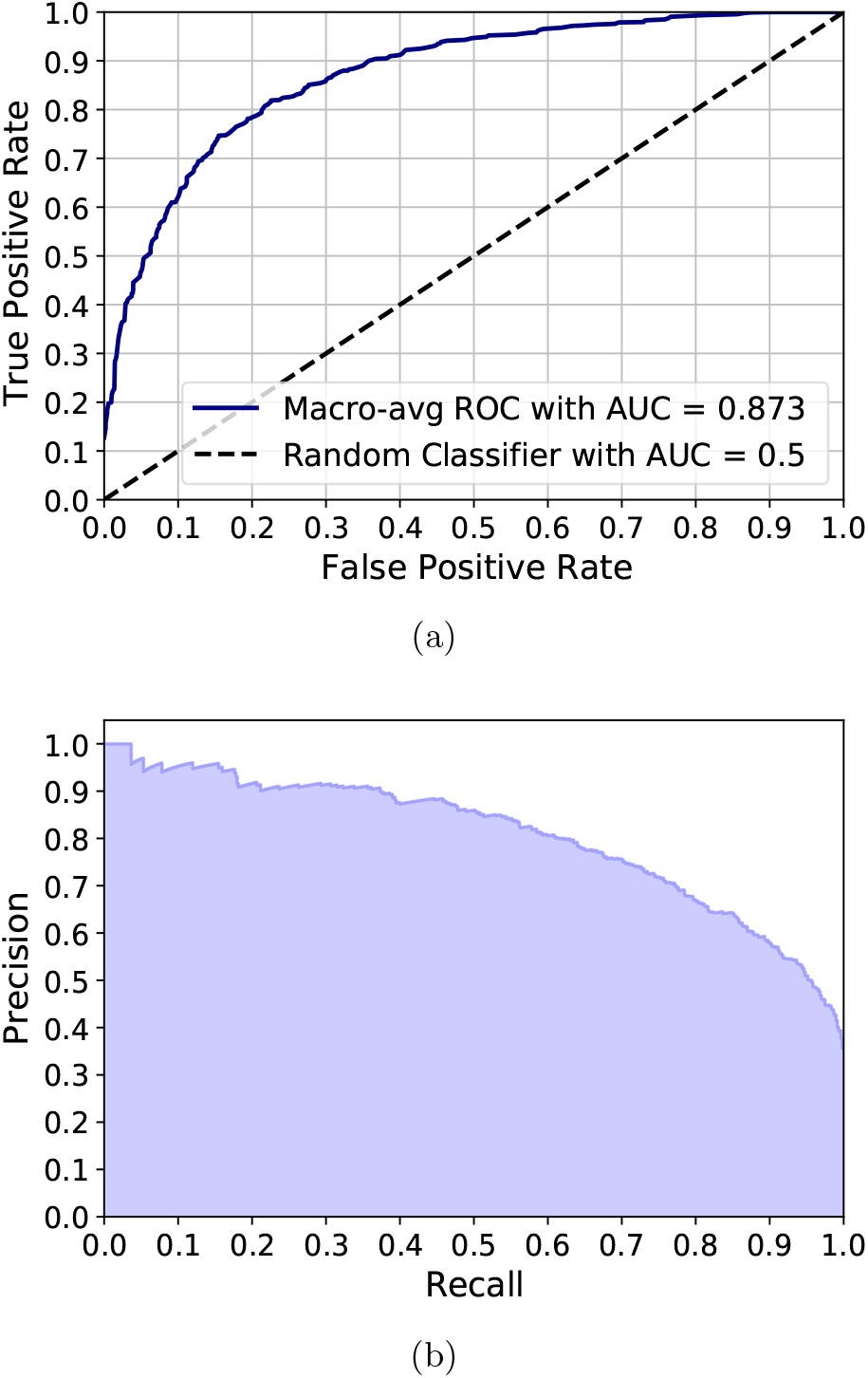
(a) ROC curve to summarize the trade-off between the true-positive rate and false-positive rate, and (b) Precision-recall curve to summarize the trade-off between the true-positive rate and the positive predictive value.

Table 5 shows the state-of-the-art comparison of our proposed pipeline with recent works, where AlexNet [33] and ResNet-101 [34] were implemented in [35] for the SLC. The proposed framework produces the best classification of the skin lesion, as shown in Table 5. Our pipeline produces the best results concerning the recall and precision beating the state-of-the-art works in [36] by a 12.0 % margin, and in [34] by a 5.0 % respectively. Our method beats method in [34] by the margin of 39.0 %, and 5.0 % in terms of the recall and precision respectively, although the AUC is same in both the methods.

**Table 5:** The state-of-the-art comparison with proposed framework, which were trained, validated, and tested on the ISIC-2017 dataset.

| Authors | Year | Recall | Precision | F1-Score | AUC |
| --- | --- | --- | --- | --- | --- |
| AlexNet [33] | 2018 | 0.34 | 0.65 | - | 0.86 |
| ResNet-101 [34] | 2018 | 0.34 | 0.71 | - | 0.87 |
| Method-1* [36] | 2018 | 0.61 | - | - | 0.84 |
| Method-2<sup>†</sup> [37] | 2018 | 0.57 | 0.68 | - | 0.85 |
| GR<sup>‡</sup> [38] | 2019 | - | - | - | 0.78 |
| **Our Proposed** | **2020** | 0.73 | 0.76 | 0.73 | 0.87 |

\*ResNet50 + RAPooling + RankOpt [36]
<sup>†</sup>ResNet50 + Eigen Decomposition + SVM [37]<sup>‡</sup>Gabor Wavelet-based CNN [38]

## 4 Conclusion

In this article, an automatic and robust framework for melanoma recognition has been proposed and implemented. The potentiality of the proposed framework has been validated via several comprehensive experiments. The scarcity of using fewer manually annotated dermoscopic images to build a generic framework was overcome by employing a transfer learning, previously trained weights, and geometric augmentation. Additional tuning the hyper-parameters and the more related augmentations may yield better recognition results for the SLC. The proposed framework will be tested on other different datasets of dermoscopic images for the SLC in the future. The proposed pipeline will be employed in other domains for recognition to verify the adaptability and generality.

## Data Availability

We use a publicly available ISIC dataset, which is free for the researchers.

